# Genetic associations with human longevity are enriched for oncogenic genes

**DOI:** 10.1101/2024.07.30.24311226

**Authors:** Junyoung Park, Andrés Peña-Tauber, Lia Talozzi, Michael D. Greicius, Yann Le Guen

**Author notes:** **Corresponding authors:** Junyoung Park, Stanford Neuroscience Health Center 290 Jane Stanford Way, Stanford, CA 94305-5090. These authors contributed equally (Michael D. Greicius, Yann Le Guen).

## Abstract

Human lifespan is shaped by both genetic and environmental exposures and their interaction. To enable precision health, it is essential to understand how genetic variants contribute to earlier death or prolonged survival. In this study, we tested the association of common genetic variants and the burden of rare non-synonymous variants in a survival analysis, using age-at-death (N = 35,551, median [min, max] = 72.4 [40.9, 85.2]), and last-known-age (N = 358,282, median [min, max] = 71.9 [52.6, 88.7]), in European ancestry participants of the UK Biobank. The associations we identified seemed predominantly driven by cancer, likely due to the age range of the cohort. Common variant analysis highlighted three longevity-associated loci: *APOE, ZSCAN23*, and *MUC5B*. We identified six genes whose burden of loss-of-function variants is significantly associated with reduced lifespan: *TET2*, *ATM*, *BRCA2, CKMT1B*, *BRCA1* and *ASXL1*. Additionally, in eight genes, the burden of pathogenic missense variants was associated with reduced lifespan: *DNMT3A, SF3B1, CHL1*, *TET2, PTEN, SOX21, TP53* and *SRSF2*. Most of these genes have previously been linked to oncogenic-related pathways and some are linked to and are known to harbor somatic variants that predispose to clonal hematopoiesis. A direction-agnostic (SKAT-O) approach additionally identified significant associations with *C1orf52, TERT, IDH2,* and *RLIM*, highlighting a link between telomerase function and longevity as well as identifying additional oncogenic genes.

Our results emphasize the importance of understanding genetic factors driving the most prevalent causes of mortality at a population level, highlighting the potential of early genetic testing to identify germline and somatic variants increasing one’s susceptibility to cancer and/or early death.

## Introduction

Longevity is a complex trait influenced by both genetic and environmental factors and their interactions [1]. According to previous studies, genetics accounts for as much as 40% of the heritability of longevity [2–4]. Identifying the genetic variants that contribute to earlier death or prolonged survival can highlight key biological pathways linked to lifespan and inform genetic testing for general health and screening and enabling precision health. Previous genome-wide association studies (GWAS) have identified over 20 associated loci including *APOE* [5, 6], *CHRNA3/5* [7], *HLA-DQA1* and *LPA* [8]. Recently, a burden analysis of protein-truncating variants from whole-exome sequencing (WES) data identified four additional genes (*BRCA2*, *BRCA1*, *ATM*, and *TET2*) linked to reduced lifespan [9]. However, most previous research on lifespan genetics has predominantly used proxy data, such as parents’ age at death, due to a lack of proband lifespan data. While proxy-based GWAS have been necessary for large cohorts of primarily middle-aged individuals with limited mortality data, this approach restricts the accuracy and scope of findings, as it may fail to comprehensively capture the genetic influences that directly impact an individual’s lifespan [10]. On the other hand, some studies have employed logistic regression models on cases of extreme longevity and younger controls [11–13]. This approach may offer new insights by focusing on exceptionally long-lived individuals, yet they can be limited and costly. Moreover, replication of borderline significant variants remains an issue due to varying case definitions across studies, with some defining cases as individuals who survive to ages beyond 90 or 100 years or using the 90th or 99th survival percentiles as the age cutoff.

In this study, we carried out a genetic analysis of direct mortality data in the UK Biobank, the genetic database with the largest number of reported deaths (35,551 subjects) and aged individuals (344,237 subjects over 60 years old). To assess the association of genetic variants with longevity in a survival analysis, we performed GWAS of common variants imputed from microarray data as well as burden/sequence kernel association test-optimized (SKAT-O) association of rare non-synonymous variants from WES data.

## Results

### Genome-wide association analyses in imputed array data

Our GWAS assessed 6,127,227 common variants (minor allele frequency (MAF) ≥ 1%) using Martingale residuals on 393,833 individuals including 35,551 deceased subjects (mean age at death: 71.2 years) and 358,282 living subjects (mean current age: 70.7) from UKB (Supplementary Table 1) [14]. Two loci reached genome-wide significance (GWS) (𝑝 < 5.0 × 10^−8^) on chromosomes 19 and 6 (Figure 1A). On chromosome 19, rs429358 was the lead variant at the *APOE* locus (β = 0.013, *p* = 6.4 × 10^-47^, MAF = 15.6%). We tested whether the presence of *APOE*-*ε*4 was enriched in certain primary causes of death. Among the top four causes of death, each representing over 5% of total deaths (Figure 1B), only those due to ‘Diseases of the circulatory system’ (Chi-square 𝑝=1.6× 10^−16^) and ‘Diseases of the nervous system’ (𝑝=1.1× 10^−71^) showed a significant enrichment in the proportion of *ε*4 carriers compared to the prevalence of *ε*4 carriers among all subjects (Figure 1C). In the chromosome 6 locus, overlapping *ZSCAN23*, two variants were GWS: rs6902687, located 2.2 kb upstream of the transcription start site (TSS), and rs13190937 situated in the 5’ untranslated region (UTR) (rs6902687_C: β = 0.004, *p* = 1.5 × 10^-8^, MAF = 36.6%; rs13190937_A: = 0.004, p = *p*= 1.5 × 10^-8^, MAF = 36.6%, Figure 1D). To explore a potential regulatory function for variants at the *ZSCAN23* locus, we investigated whether the lead SNPs were expression quantitative trait loci (eQTLs) in the Genotype-Tissue Expression Project (GTEx) v8 database. rs13190937 was significantly associated with increased *ZSCAN23* expression in pancreatic tissue and the longevity GWAS signal colocalized with the *ZSCAN23* expression quantitative trait loci (eQTL) (posterior probability of colocalization (PP4) = 0.94; Figure 1E). Phenome-wide association study analysis (PheWAS) using PheWeb [15] shows that the main associations of rs13190937 are with celiac disease and intestinal malabsorption (𝑝 =1.8 × 10^−57^) (Supplementary Figure 1).

**Figure 1.**
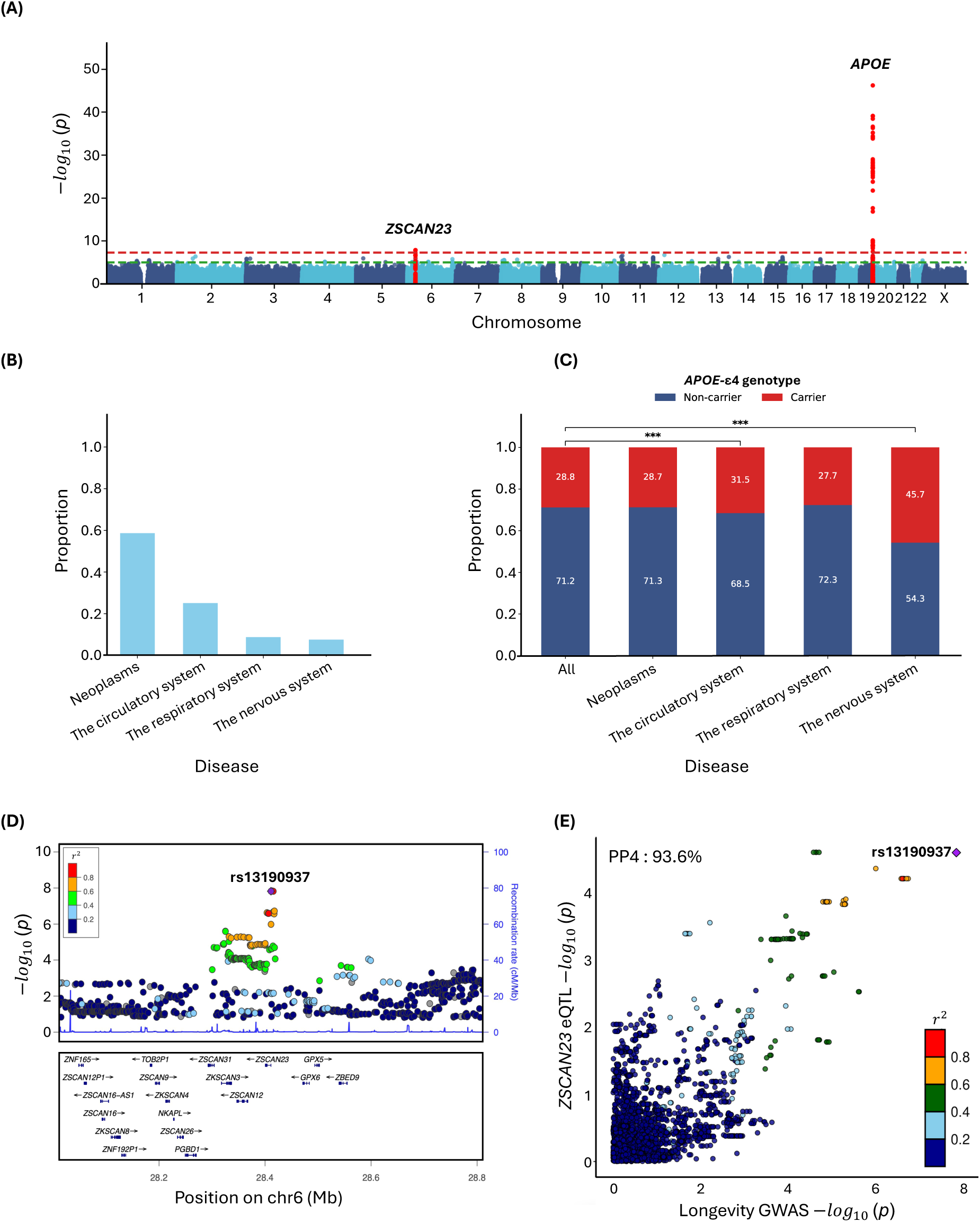
Common variant GWAS of longevity. (A) Manhattan plot. (B) The proportion of cause of death for the top 4 categories, each accounting for more than 5% of total deaths) (C) Association of causes of death with *APOE-ε*4 genotype. (D) Locuszoom and (E) colocalization plots at the *ZSCAN23* locus, colocalized with *ZSCAN23* eQTL in pancreatic tissue in GTEx. PP4: posterior probability of colocalization.

In sex-stratified GWAS (180,970 males and 212,863 females), the *APOE* locus was again linked to longevity in both males and females (Supplementary Table 1 and Supplementary Figure 2A and B). In males, we observed an additional GWS association for rs35705950_T located between *MUC5AC* and *MUC5B* on chromosome 11 (β = 0.01, *p* = 2.1 × 10^-8^, MAF = 11.2%) (Supplementary Figure 2C), while no additional association was found in females. This variant was notably linked to increased *MUC5B* expression in lung tissue with the longevity GWAS signal aligning with a *MUC5B* eQTL (PP4 = 0.99; Supplementary Figure 2D). We also confirmed through PheWAS that rs35705950 is associated with a diagnosis of pulmonary fibrosis (𝑝=4.4× 10^−13^) and “Other interstitial pulmonary diseases with fibrosis” listed as primary cause of death (𝑝=1.7× 10^−5^), and Illness of the father “Lung cancer” (𝑝=2.1× 10^−4^), but not with the mother’s (𝑝=0.07) (Supplementary Figure 2E).

### Gene-based rare variant association analyses in whole-exome data

Among 26,230,624 variants with MAF < 1%, 1,830,070 variants (17,174 genes) were annotated as loss-of-function (LoF) or missense variants. Of these, 628,362 were predicted LoF variants (17,071 genes with a median of 28 variants per gene), 985,950 were missense variants predicted as damaging by AlphaMissense (15,891 genes with a median of 47 variants per gene), and 349,791 were missense variants predicted as damaging by rare exome variant ensemble learner (REVEL) (12,219 genes with a median of 12 variants per gene). Of variants classified by each, 23.7% of AlphaMissense and 66.9% of REVEL variants were also pathogenic by the other classifier.

We identified six genes whose burden of LoF variants is significantly associated with reduced lifespan: *TET2* (𝑝=3.8× 10^−30^), *ATM* (𝑝=6.0× 10^−10^), *BRCA2* (𝑝=1.3× 10^−34^), *CKMT1B* (𝑝=4.5× 10^−7^), *BRCA1* (𝑝=4.9× 10^−12^) and *ASXL1* (𝑝=2.3× 10^−44^) (Figure 2A and Table1). All of these but *CKMT1B* also showed gene-wide significance in a direction-agnostic (SKAT-O) approach (Supplementary Figure 3A). Additionally, in eight genes, the burden of missense variants predicted as pathogenic by AlphaMissense was associated with reduced lifespan: *DNMT3A* (𝑝=1.4× 10^−9^), *SF3B1* (𝑝=6.7× 10^−12^), *CHL1* (𝑝=5.0× 10^−7^), *TET2* (𝑝=4.2× 10^−7^), *PTEN* (𝑝=1.0× 10^−8^), *SOX21* (𝑝=2.9× 10^−8^), *TP53* (𝑝=3.1× 10^−15^) and *SRSF2* (𝑝=9.8× 10^−89^) (Figure 2B). Lastly, three genes showed gene-wide significance for burden of missense variants predicted by REVEL: *DNMT3A* (𝑝 =6.6 × 10^−9^), *PTEN* (𝑝=6.6× 10^−8^), and *TP53* (𝑝=6.2× 10^−9^) (Supplementary Figure 4 and Supplementary Table 2). SKAT-O identified additional associations with pathogenic missense variants predicted by AlphaMissense in *C1orf52* (𝑝=2.1× 10^−7^), *IDH2* (𝑝=5.3 × 10^−39^) and *RLIM* (𝑝=3.7× 10^−9^) (Supplementary Figure 3B), and by REVEL in *TERT* (𝑝=8.1× 10^−10^) (Supplementary Figure 3C and Supplementary Table 2).

**Figure 2.**
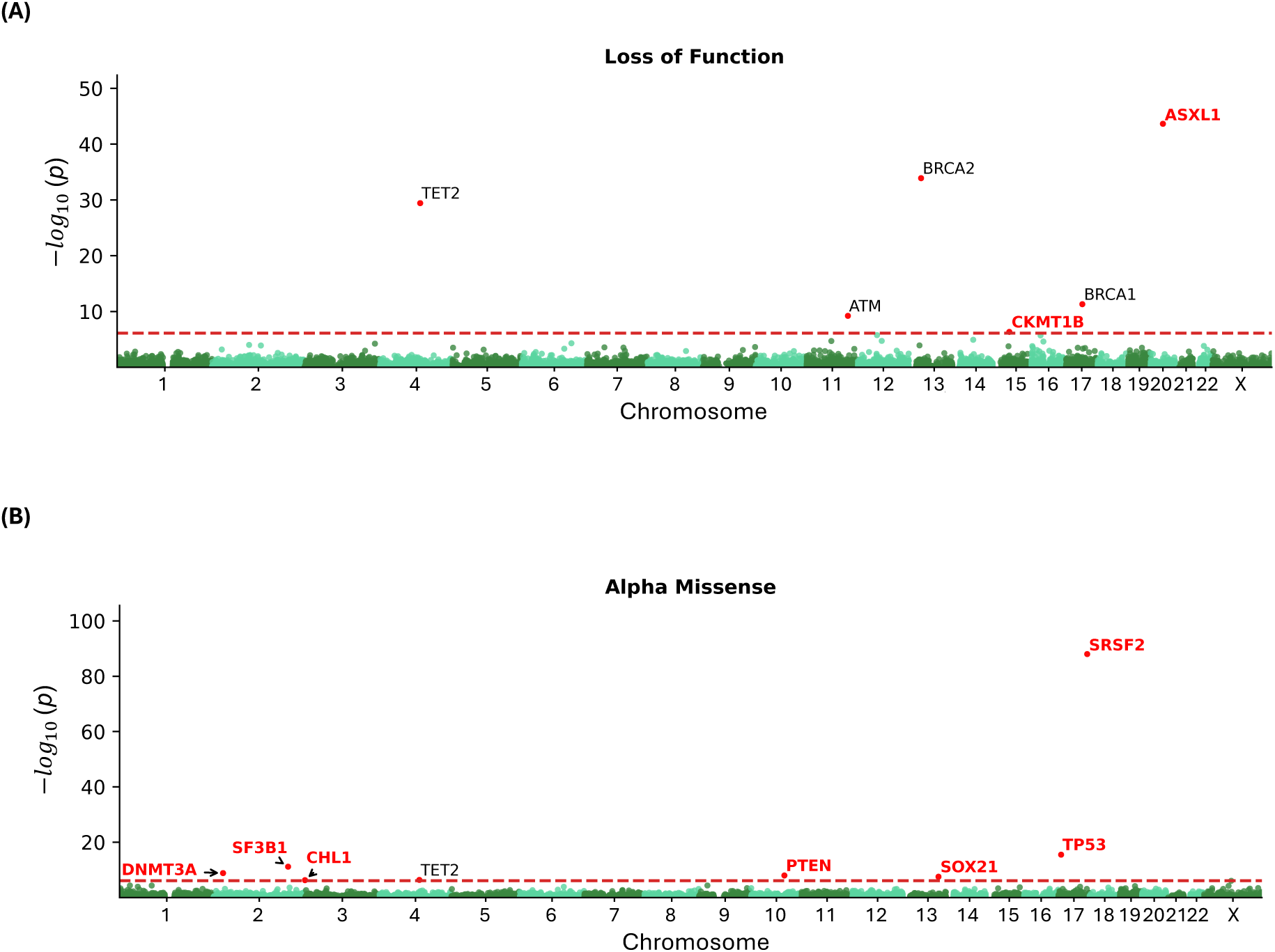
Rare variant burden association with longevity, considering loss-of-functions (A) and Alpha Missense pathogenic variants (B). Novel genes are highlighted in red.

For sex-specific gene-based analysis, an additional six genes not identified in the whole-cohort analysis showed gene-wide significance in males by either burden or SKAT-O: *CDKN1A* and *PTPRK* (LoF); *COA7* and *TG* (AlphaMissense); *NMNAT2* and *PITRM1* (REVEL) (Supplementary Figure 5A, 6A and Supplementary Table 3). In females, we identified three additional genes associated with reduced lifespan: *PORCN* (AlphaMissense); *UGT1A8* and *OLIG1* (REVEL) (Supplementary Figure 5B, 6B and Supplementary Table 4).

### Gene-burden survival analysis

For the 13 gene-wide significant genes in the burden analyses, we assessed the association of variant carrier status with lifespan using Cox proportional hazards regression. Carriers of LoF variants in six genes were associated with decreased survival compared to non-carriers: *CKMT1B* (HR=3.9, 𝑝 =2.1 × 10^−6^), *ASXL1* (HR=2.2, 𝑝 =3.8 × 10^−26^) (Figure 3A), *TET2* (HR=1.7, 𝑝 = 2.7× 10^−18^), *ATM* (HR=1.7, 𝑝=2.5× 10^−10^), *BRCA2* (HR=2.4, 𝑝=1.1× 10^−40^), and *BRCA1* (HR =2.2, 𝑝=1.0 × 10^−12^) (Supplementary Figure 7A). Similarly, carriers of AlphaMissense-predicted pathogenic variants exhibited significantly earlier mortality compared to non-carriers on the following genes: *DNMT3A* (HR=1.4, 𝑝=2.0× 10^−7^), *SF3B1* (HR=2.1, 𝑝=2.0× 10^−8^), *CHL1* (HR=1.3, 𝑝=3.4× 10^−7^), *PTEN* (HR=4.2, 𝑝=3.4× 10^−10^), *SOX21* (HR=1.9, 𝑝=5.2× 10^−8^), *TP53* (HR=3.7, 𝑝=1.7× 10^−13^), *SRSF2* (HR=5.0, 𝑝=2.4×10^−52^) (Figure 3B) and *TET2* (HR=1.5, 𝑝=3.8× 10^−6^) (Supplementary Figure 7B). Carriers of pathogenic variants predicted by REVEL showed similar trends: *DNMT3A* (HR=1.5, 𝑝 =1.9 × 10^−6^), *PTEN* (HR=5.3, 𝑝 =1.3 × 10^−10^), and *TP53* (HR=2.4, 𝑝 =6.6 × 10^−8^) (Supplementary Figure 7C).

**Figure 3.**
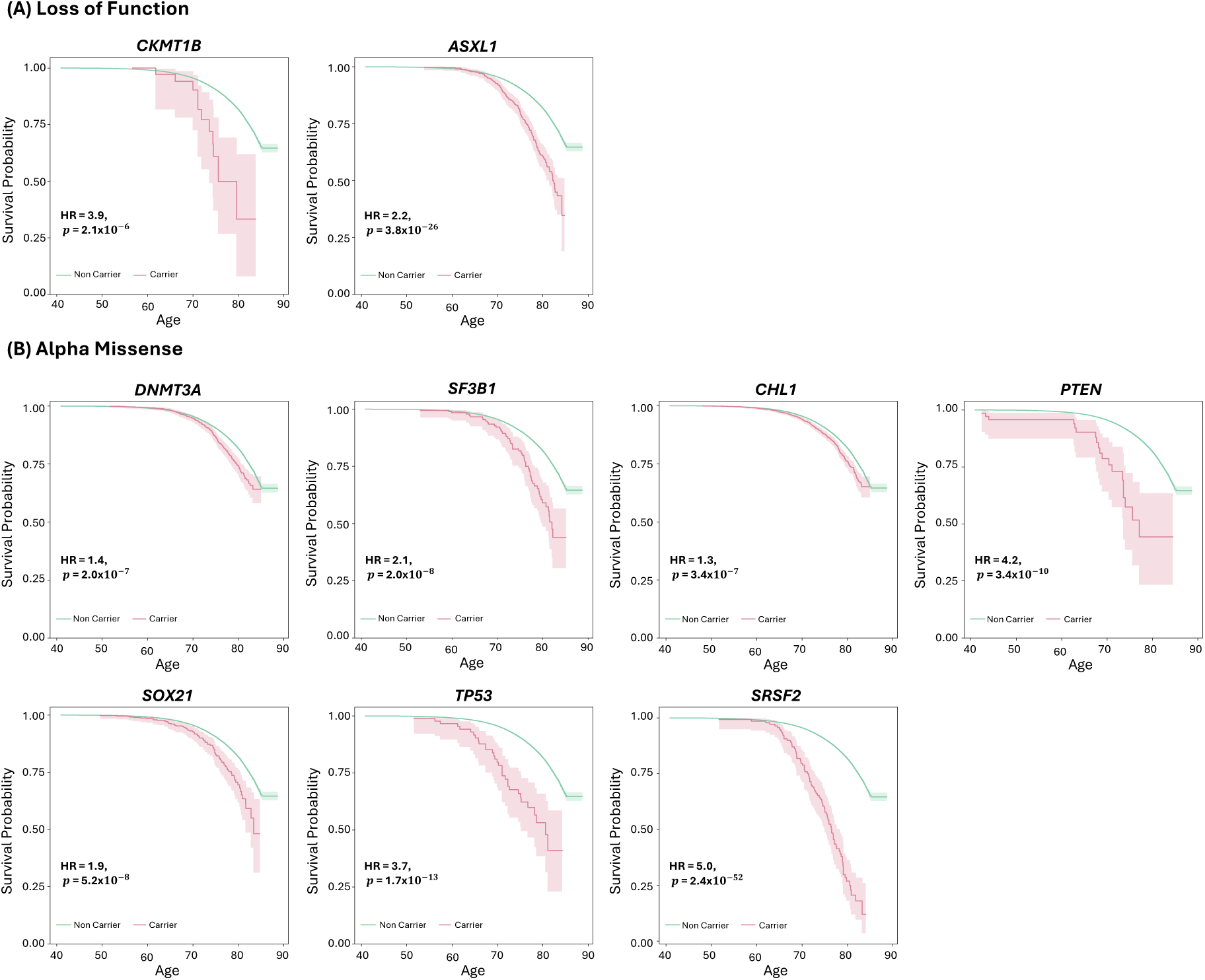
Survival curves comparing carriers and non-carriers of variants on genes with a significant burden of loss-of-function (A) and AlphaMissense pathogenic (B) variants.

To explore the contribution of individual rare variants to mortality in each gene-wide significant gene in the burden and SKAT-O tests, we conducted Cox proportional hazards regression for each variant with a minor allele count (MAC) of three or more (Table 2). In total, 587 variants including LoF, AlphaMissense and REVEL variants were examined. After applying a Bonferroni correction for multiple testing, setting the significance threshold at 8.5× 10^−5^ (0.05/587), we identified significant associations with reduced lifespan for four LoF variants: rs370735654 in *TET2* (MAC=17, HR=7.0, 𝑝=5.6× 10^−9^), rs587779834 in *ATM* (MAC=113, HR=2.5, 𝑝=2.7× 10^−5^), rs80359520 in *BRCA2* (MAC=10, HR=6.1, 𝑝 =1.8×10^−6^), rs750318549 in *ASXL1* (MAC=201, HR=2.5, 𝑝=2.3× 10^−15^). Additionally, significant associations with AlphaMissense variants were noted in six genes, impacting lifespan: rs769009649 in *C1orf52* (MAC=62, HR=3.2, 𝑝 =7.4 × 10^−7^), rs147001633 in *DNMT3A* (MAC=269, HR=1.8, 𝑝 =3.7 × 10^−5^), rs377023736 in *SF3B1* (MAC=12, HR=6.0, 𝑝 =8.5 × 10^−8^), rs116421102 in *CHL1* (MAC=1,842, HR=1.3, 𝑝=2.5× 10^−5^), rs121913502 in *IDH2* (MAC=45, HR=5.7, 𝑝=1.0× 10^−20^), rs11540652 in *TP53* (MAC=5, HR=10.0, 𝑝=6.6× 10^−8^) and rs751713049 in *SRSF2* (MAC=51, HR=5.8, 𝑝 =1.9 × 10^−26^). For missense variants predicted by REVEL, rs1043358053 in *TERT* (MAC=5, HR=11.9, 𝑝 =7.4 × 10^−5^) and rs11540652 in *TP53* were significantly linked to reduced lifespan (Supplementary Table 5).

### Phenome-wide association studies

For the nine novel longevity genes identified in the burden test (*CKMT1B, ASXL1, DNMT3A, SF3B1, CHL1*, *PTEN, SOX21, TP53* and *SRSF2*), we examined the burden of LoF or pathogenic missense variants through PheWASs across 1,670 UKB phenotypes including disease occurrences derived from electronic health record, self-reported family history, and physical measures (Supplementary Figure 8). The burden of LoF variants in *ASXL1* and AlphaMissense variants in *DNMT3A, SF3B1, PTEN*, *TP53* and *SRSF2* were strongly linked to an increased risk of leukemia: acute myeloid leukemia (*ASXL1*: Odds Ratio (OR)=1.05; 𝑝=8.6× 10^−170^; *DNMT3A*: OR=1.03, 𝑝=2.1× 10^−150^; *SRSF2*: OR=1.3, 𝑝=1.2× 10^−195^; *TP53*: OR=1.05, 𝑝=4.7× 10^−35^), monocytic leukemia (*DNMT3A*: OR=1.01, 𝑝=2.5 × 10^-9^), chronic lymphoid leukemia (*SF3B1*: OR=1.07, 𝑝=4.1× 10^−68^) and acute lymphoid leukemia (*PTEN*: OR=1.01, 𝑝 =2.1 × 10^−14^). Additionally, the burden of LoF in *CKMT1B* was associated with hypopharynx cancer (OR=1.03, 𝑝 =3.9 × 10^−26^), vertiginous syndromes (OR=1.03, 𝑝=3.0× 10^−17^) and salivary glands cancer (OR=1.03; 𝑝=3.2× 10^−12^). *SOX21* burden was associated with increased acne (OR=1.01, 𝑝=6.9× 10^−7^) and spinocerebellar disease (OR=1.01, 𝑝=2.3× 10^−6^).

### Somatic mutation and clonal hematopoiesis of indeterminate potential

We computed the variant allelic fraction (VAF) per carrier for each variant included in the analysis. Generally, germline variants have a mean VAF close to 50%, while somatic variants’ mean VAF will be lower [16]. Thus, when an association is linked to clonal hematopoiesis of indeterminate potential (CHIP), we expect the distribution of VAF to be left-shifted compared to a normal distribution centered at VAF = 50%. Considering LoF variants, *TET2* (mean VAF across variants [95% bootstrap confidence interval for the mean VAF] = 0.33 [0.31,0.34]) and *ASXL1* (mean VAF =0.32 [0.31,0.33]) burden test associations are supported by variants with a left-shifted VAF distribution (Supplementary Table 6, Supplementary Figure 9A). Similarly, considering pathogenic Alpha Missense variants, in *DNMT3A* (mean VAF= 0.24 [0.23-0.24]), *TET2* (mean VAF=0.36 [0.34,0.38]), *TP53* (mean VAF=0.28 [0.24,0.34]), *SRSF2* (mean VAF=0.30 [0.28,0.31]), *SF3B1* (mean VAF=0.31 [0.26,0.37]) and CHL1 (mean VAF=0.37[0.28-0.45]) are also left-shifted and the observed associations may be linked to CHIP (Supplementary Table 5, Supplementary Figure 9B).

## Methods

### Study participants

The UKB is a large population-based longitudinal cohort study with recruitment from 2006 to 2010 in the United Kingdom [17]. In total, 502,664 participants aged 40-69 years were recruited and underwent extensive phenotyping including health and demographic questionnaires, clinic measurements, and blood draw at one of 22 assessment centers, of whom 468,541 subjects have been genotyped by both SNP array and WES.

We restricted our analysis to 393,833 individuals who self-reported their ethnic background as ‘white British’ and were categorized as European ancestry based on genetic ethnic grouping (Field: 21000). Among them, 35,551 subjects were reported deceased, and their ages at death were recorded from the UK Death Registry (Field: 40007). For the other 358,282 subjects without death records, we assumed they were still alive by the latest censoring date (November 30, 2023). We determined their last known ages by subtracting their year and month of birth (Field: 33) from the censoring date.

### SNP array genotyping and QC

A total of 488,000 UKB participants were genotyped using one of two closely related Affymetrix microarrays (UKB Axiom Array or UK BiLEVE Axiom Array) for ∼ 820,000 variants. Quality control (QC), phasing, and imputation were performed as described previously [18]. Briefly, the genotyped dataset was phased and imputed into UK10K, 1000 Genomes Project phase 3, and Haplotype Reference Consortium reference panels, resulting in approximately 97 million variants. Additionally, we removed SNPs with imputation quality score < 0.3, genotype missing rate > 0.05, minor allele frequency (MAF) < 1%, and Hardy-Weinberg equilibrium 𝑝 < 1.0 × 10^−6^.

### Genome-wide association studies

We performed linear regression models using PLINK v2.0 [19] to test the association of common variants (MAF ≥ 1%) with longevity for the entire cohort, as well as stratified by sex. For all three analyses, we used Martingale residuals calculated using the Cox proportional hazards model as the outcome variable. The procedure for calculating Martingale residuals was as follows. First, a Cox proportional hazards model [20] was fitted without genotype,

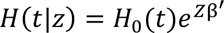

where 𝐻_0_(𝑡) is the baseline hazard function at time point 𝑡 given the last-known age and dead/alive status, Z = [Z_1_, …, Z_𝑘_] is a covariate matrix, and β = [β_1_, …, β_𝑘_] is a coefficient matrix for Z. Here, we included sex and the first five principal components (PC) as covariates, but for sex-specific analyses, sex was excluded. Then, Martingale residuals were calculated as:

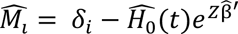

where 𝛿_𝑖_ is the dead/alive status (0=alive, 1=dead) of the 𝑖th subject and β< is the estimated coefficient matrix. We adapted the *coxph* function from the *survival* (v.3.6.4) R package [21] to compute the Martingale residuals. Genome-wide significance threshold was set at the standard GWAS level (𝑝=5.0 × 10^−8^). We used *LocusZoom* [22] to generate regional plots and Python v.3.7 to create Manhattan plots.

### Gene expression and colocalization analysis

To evaluate the effect of the significant loci identified in our GWAS, we examined expression quantitative trait loci (eQTLs) across 49 tissues having at least 73 samples from the Genotype-Tissue Expression Project (GTEx) version 8 [23]. Bayesian colocalization analysis was employed using the *COLOC* package (v.5.2.3) [24] in R and the posterior probability of colocalization (PP4) was calculated between GWAS findings and eQTL associations within a 1 megabase (Mb) window. Additionally, colocalization was visualized using the *locuscompareR* package [25].

### Whole-exome sequencing and QC

Whole exome sequencing (WES) data was available for 469,835 UKB participants. The dataset was generated by the Regeneron Genetics Center [26]. Details about the production and QC for the WES data are described previously [26]. We restricted the WES analysis to rare variants (MAF < 1%).

### Rare variant annotation

Rare variants in WES data were annotated using Variant Effect Predictor (v. 112) provided by Ensembl [27]. We defined LoF variants as those with predicted consequences: splice acceptor, splice donor, stop gained, frameshift, start loss, stop loss, transcript ablation, feature elongation, or feature truncation. Missense variants were annotated using AlphaMissense [28] and REVEL [29] plugins and included if they had an AlphaMissense score ≥ 0.7 or REVEL score ≥ 0.75. All annotation was conducted based on GRCh38 genome coordinates.

### Gene-based rare variant association studies

For testing groups of rare variants, genotype matrices were first transformed into a binary variable describing whether samples carry a variant of a given class as follows:

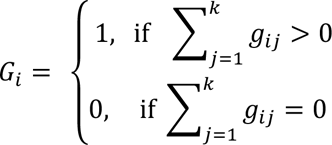

Where 𝑔_𝑖j_ is the minor allele count observed for subject 𝑖 at variant 𝑗 in the gene and 𝑘 is the number of variants in the gene. We carried out two gene-based tests: Burden test and sequence kernel association test-optimized (SKAT-O) [30]. The burden test is a mean-based test that assumes the same direction of effects for all variants within a gene. On the other hand, SKAT-O employs a weighted average of the burden test and SKAT [31], the latter a variance-based test that does not lose power when variants have opposing directions of effect.

Association tests were performed for each gene and rare variant class, including separately LoF variants, missense variants with an AlphaMissense score ≥ 0.7, and missense variants with a REVEL score ≥ 0.75, using Martingale residuals as the phenotype as in the common variant analyses. We excluded genes with fewer than 10 variant carriers to ensure the reliability of our analyses. A gene-wide significance threshold was established at 𝑝=7.4× 10^−7^based on the Bonferroni method accounting for the number of genes, variant classes, and statistical methods. Gene-based analyses were carried out using the *SKAT* package (v.2.2.5) in R.

To characterize the impacts of gene burden in significant genes, we compared lifespan survival depending on gene burden using Kaplan–Meier survival curves, log-rank tests, and Cox proportional hazard regression analyses. Additionally, we performed Cox proportional hazards regression to assess the effect of each rare variant in a gene. The *survival* (v.3.6.4) package in R was utilized for the survival analysis.

### Phenome-wide association studies

For gene-wide significant genes, we conducted phenome-wide association studies (PheWAS) of variant carrier status across 1,670 phenotypes in the UKB derived from binary, categorical, and continuous traits. Phenotypes included the International Classification of Disease 10 (ICD-10) codes, family history (e.g. father’s illness, father’s age at death), blood count (e.g. white blood cell count), blood biochemistry (e.g. Glucose levels), infectious diseases (e.g. pp 52 antigen for Human Cytomegalovirus), physical measures (e.g. BMI), cognitive test (e.g. pairs matching) and brain measurements (e.g. subcortical volume of hippocampus). For ICD-10 codes, we excluded phenotypes from the following ICD-10 chapters: ‘Injuries, poisonings, and certain other consequences of external causes’ (Chapter XIX), ‘External causes of morbidity and mortality’ (Chapter XX), ‘Factors influencing health status and contacts with health services’ (Chapter XXI), and ‘Codes for special purposes’ (Chapter XXII). The ICD-10 codes were then converted into Phecodes (v.1.2) [32] which combine correlated ICD codes into a distinct code and improve alignment with diseases commonly used in clinical practice.

For binary traits, we removed phenotypes with fewer than 100 cases, and for continuous traits, those with fewer than 100 participants were excluded. Depending on the phenotype, we employed various regression models including binary logistic regression, ordinal logistic regression, multinomial logistic regression, and linear regression. All analyses included age, sex, and first five PCs as covariates. Phenome-wide significance threshold was set at 𝑝=2.9 × 10^−5^ based on the number of phenotypes.

### Variant allelic fraction

To investigate whether some gene-level associations are enriched for somatic variants, we computed the variant allele frequency (VAF) for each heterozygous sample, reporting the mean VAF and VAF distribution per gene per variant class. VAF is defined as the number of reads with an alternate allele divided by the read depth at a given variant position. We also calculated the confidence interval for the mean VAF per gene using 10,000 bootstrap samples to ensure robust statistical analysis.

## Discussion

In this study, we report several known and novel findings related to genetic risks associated with longevity analyzing 393,833 European participants from the UKB. In the common variant GWAS, three independent loci associated with increased mortality risk were identified. In the gene-based analysis of rare non-synonymous variants, 17 genes had their burden/SKAT-O test associated with longevity.

Consistent with previous reports, rs429358, determining the *APOE-ε*4 allele dosage, was associated with decreased lifespan across both sexes. *APOE*-*ε*4 is well known for its associations with Alzheimer’s Disease [33] and cardiovascular disease [34]. In our dataset, the proportion of *ε*4 carriers was significantly higher for deaths caused by ‘Disease of the circulatory system’ and ‘Diseases of the nervous system’ compared to the general prevalence of ε4 carriers, which could explain the effect of *ε*4 on longevity. Examining the subcategories of these ICD-10 chapters, ‘Disease of the circulatory system’ includes cardiovascular disease (I51.6), while ‘Diseases of the nervous system’ covers Alzheimer’s disease (G30). We also identified a GWS association at the *ZSCAN23* locus, which had not been previously reported. Our colocalization analysis revealed that the longevity-associated signal colocalizes with a *ZSCAN23* eQTL in pancreatic tissue with increased expression observed in minor allele carriers. Although the role of *ZSCAN23* remains unclear, recent studies have linked its expression to pancreatic tumors, supporting our colocalization findings [35]. For sex-specific GWAS, a GWS association specific to males was found between *MUC5AC* and *MUC5B*, which highly colocalizes with a *MUC5B* eQTL in lung tissue and many studies have linked this variant to pulmonary disease like idiopathic pulmonary fibrosis [36, 37] and COVID-19 [38, 39]. Previously reported SNP associations with longevity were concordant in our dataset but none of these passed the GWAS suggestive threshold (𝑝=1.0× 10^−5^) except for those at the *APOE* locus. This phenomenon likely resulted from previous studies relying on proxy data such as parental age at death, which may capture a different set of genetic factors than direct proband mortality data.

In our gene-based rare variant analysis, 17 genes achieved gene-wide significance (𝑝 <7.4× 10^−7^) in either the burden or SKAT-O test. Four of these, *TET2*, *ATM*, *BRCA2*, and *BRCA1*, were reported in a previous rare-variant analysis of longevity in UKB [9]. We identified 13 novel genes associated with longevity—*CKMT1B*, *ASXL1*, *DNMT3A*, *SF3B1*, *PTEN*, *SOX21*, *TP53*, *SRSF2*, *C1orf52*, *CHL1*, *IDH2*, *RLIM*, and *TERT*— when assessing variants causing genetic LoF or missense variants classified as pathogenic by REVEL or AlphaMissense. Of note, LoF and missense variant analyses identified mostly separate genes with only one overlap (*TET2*). This supports the use of both categories in rare variant analyses and may indicate that missense variants as classified by AlphaMissense capture a wider range of variation missed when only assessing LoF variants, which are generally interpreted as resulting in haploinsufficiency. Importantly, missense variants may lead to increased or decreased protein function. In our analyses, *IDH2* was not gene-wide significant with the burden test (𝑝=1.9× 10^−4^, Table 1) but was highly significant with SKAT-O (𝑝=5.3× 10^−39^, Table 1). Since SKAT-O does not lose power when variants have differing directions of effect, this suggests that different mutations in *IDH2* can lead to either increased or decreased longevity. These results underline the gain in information achieved when studying rare missense variants as well as LoF using appropriate statistical techniques.

**Table 1.** Significant genes for rare variants association with burden and SKAT-O tests (𝒑<7.4× 𝟏𝟎^−7^). Gene names in bold font represent novel associations.

| Variant Class | Chr | Gene | # of variants | # of carriers | Burden<br>*p*-value | SKAT-O<br>*p*-value |
| --- | --- | --- | --- | --- | --- | --- |
| LoF | 4 | *TET2* | 243 | 563 | $3.8 \times 10^{-30}$ | $3.4 \times 10^{-54}$ |
| | 11 | *ATM* | 247 | 1,170 | $6.0 \times 10^{-10}$ | $4.8 \times 10^{-11}$ |
| | 13 | *BRCA2* | 245 | 1,271 | $1.3 \times 10^{-34}$ | $1.2 \times 10^{-42}$ |
| | 15 | ***CKMT1B*** | 15 | 40 | $4.5 \times 10^{-7}$ | $1.8 \times 10^{-6}$ |
| | 17 | *BRCA1* | 120 | 456 | $4.9 \times 10^{-12}$ | $1.3 \times 10^{-11}$ |
| | 20 | ***ASXL1*** | 72 | 533 | $2.3 \times 10^{-44}$ | $2.9 \times 10^{-46}$ |
| Alpha Missense | 1 | ***Clorf52*** | 23 | 175 | $4.5 \times 10^{-5}$ | $2.1 \times 10^{-7}$ |
| | 2 | ***DNMT3A*** | 167 | 1,229 | $1.4 \times 10^{-9}$ | $1.8 \times 10^{-10}$ |
| | 2 | ***SF3B1*** | 64 | 195 | $6.7 \times 10^{-12}$ | $2.5 \times 10^{-16}$ |
| | 3 | ***CHL1*** | 33 | 3,666 | $5.0 \times 10^{-7}$ | $1.1 \times 10^{-6}$ |
| | 4 | *TET2* | 159 | 826 | $4.2 \times 10^{-7}$ | $7.4 \times 10^{-7}$ |
| | 10 | ***PTEN*** | 50 | 71 | $1.0 \times 10^{-8}$ | $3.9 \times 10^{-11}$ |
| | 13 | ***SOX21*** | 52 | 463 | $2.9 \times 10^{-8}$ | $6.7 \times 10^{-9}$ |
| | 15 | ***IDH2*** | 89 | 349 | $1.9 \times 10^{-4}$ | $5.3 \times 10^{-39}$ |
| | 17 | ***TP53*** | 35 | 90 | $3.1 \times 10^{-15}$ | $2.5 \times 10^{-16}$ |
| | 17 | ***SRSF2*** | 14 | 141 | $9.8 \times 10^{-89}$ | $8.3 \times 10^{-108}$ |
| | X | ***RLIM*** | 25 | 51 | $8.8 \times 10^{-7}$ | $3.7 \times 10^{-9}$ |

LoF: Loss of Function; Chr: chromosome

**Table 2.** Lead variant association per gene among significant genes in the burden and SKAT-O tests. . Only variants with at least 3 minor alleles are reported.

| Variant Class | Chr | Gene | Variant | MA | MAC | AM | HR | *p*-value | Reported |
| --- | --- | --- | --- | --- | --- | --- | --- | --- | --- |
| LoF | 4 | *TET2* | rs370735654 | T | 17 | - | 7.0 | $5.6 \times 10^{-9}$ | - |
| | 11 | *ATM* | rs587779834 | A | 113 | - | 2.5 | $2.7 \times 10^{-5}$ | - |
| | 13 | *BRCA2* | rs80359520 | C | 10 | - | 6.1 | $1.8 \times 10^{-6}$ | Breast Cancer [66] |
| | 15 | *CKMT1B* | rs1355844751 | T | 8 | - | 4.9 | $5.9 \times 10^{-3}$ | - |
| | 17 | *BRCA1* | rs80357508 | C | 44 | - | 2.8 | $3.5 \times 10^{-3}$ | - |
| | 20 | *ASXL1* | rs750318549 | AG | 201 | - | 2.5 | $2.3 \times 10^{-15}$ | - |
| Alpha Missense | 1 | *C1orf52* | rs769009649 | A | 62 | 0.876 | 3.2 | $7.4 \times 10^{-7}$ | - |
| | 2 | *DNMT3A* | rs147001633 | T | 269 | 0.995 | 1.8 | $3.7 \times 10^{-5}$ | Leukemia [67, 68] |
| | 2 | *SF3B1* | rs377023736 | A | 12 | 0.999 | 6.0 | $8.5 \times 10^{-8}$ | - |
| | 3 | *CHL1* | rs116421102 | C | 1,842 | 0.745 | 1.3 | $2.5 \times 10^{-5}$ | - |
| | 4 | *TET2* | rs76428136 | G | 5 | 0.913 | 8.1 | $2.9 \times 10^{-4}$ | - |
| | 10 | *PTEN* | rs587782350 | T | 3 | 0.941 | 20.4 | $2.6 \times 10^{-3}$ | - |
| | 13 | *SOX21* | rs1172148601 | A | 67 | 0.856 | 2.3 | $1.7 \times 10^{-3}$ | - |
| | 15 | *IDH2* | rs121913502 | T | 45 | 0.987 | 5.7 | $1.0 \times 10^{-20}$ | Leukemia [68, 69] |
| | 17 | *TP53* | rs11540652 | T | 5 | 0.996 | 10.0 | $6.6 \times 10^{-5}$ | Gastric Cancer [70],<br>Ovarian Cancer [71] |
| | 17 | *SRSF2* | rs751713049 | T | 51 | 0.982 | 5.8 | $1.9 \times 10^{-26}$ | - |
| | X | *RLIM* | X:74592998:C:A | A | 3 | 0.970 | 3.9 | $1.3 \times 10^{-3}$ | - |

Chr: chromosome; MAC: minor allele count; AM: AlphaMissense score; HR: hazard ratio

Strikingly, most of the genes we identified carrying longevity-associated rare variants have been previously linked to cancer. *TET2, ASXL1, DNMT3A*, and *SF3B1* are all known to harbor causal leukemia variants [40–43], and somatic variants in *SRSF2* have been described in myelodysplastic syndrome [44]. *ATM, BRCA2*, and *BRCA1* mutations have been well characterized in breast, ovarian, and other cancers [45–47]. *RLIM* appears to be a regulator of estrogen-dependent transcription, an important pathway in breast cancer [48], and has been recently described as a potential tumor suppressor [49]. *PTEN* and *TP53* are well studied due to their critical role in genomic stability and are the two most mutated genes in human cancer [50]. *IDH2* is also frequently mutated in many kinds of cancer [51]. The antisense long noncoding RNA *SOX21-AS1*, but not *SOX21*, has been linked to oral, cervical, and breast cancer [52–54]. A recent study found potential for *CKMT1B* expression as a prognostic biomarker in glioma [55]; similarly, alterations in CHL1 expression have been associated with development and metastasis of many types of cancer [56]. Finally, variation in both the coding and promoter sequences of *TERT* has been associated with a variety of cancer types [57, 58]. Our PheWAS results also suggest that most of these genes are associated with cancer, specifically blood-based tumors such as myeloid leukemia. Combined with the common *ZSCAN23* locus we identified, associated with pancreatic tumors, this points to cancer being the major genetic factor currently affecting lifespan in UKB. This is consistent with a previous study of healthspan that found cancer to be the first emerging disease in over half of disease cases in UKB [59]. These results likely reflect the characteristics of the cohort, comprised of predominantly middle-aged individuals, with age-at-death ranging from 40.9 to 85.2 years and last-known ages between 52.6 and 88.7 years.

For sex-specific rare variant analyses, we identified six novel genes (*CDKN1A*, *PTPRK*, COA7, *TG*, *NMNAT2* and *PITRM1*) in males and three genes (*PORCN*, *UGT1A8* and *OLIG1*) in females. Some of these genes have been found to associate with sex-specific diseases. In one study, advanced prostate cancer patients had a higher frequency of a variant on the 3’UTR of CDKN1A [60] and the gene has received attention as a potential therapeutic target for prostate cancer [61]. *PORCN* is located on the X chromosome and mutations on it can cause Goltz-Gorlin Syndrome [62], but it has also been found to regulate a signaling pathway that controls cancer cell growth [63]. *UGT1A8* expression is altered in endometrial cancer [64] and amino acid substitutions in it may modulate estradiol metabolism leading to increased risk of breast and endometrial cancer [65].

Since UKB collected DNA from peripheral blood mononuclear cell samples, we explored whether the variants were potentially of somatic origin, picked up by WES genotyping due to CHIP. The VAF distribution of variants included in our analysis emphasizes that several associations are likely linked to CHIP, and notably include the well-established CHIP-related genes *TET2, ASLX1*, *DNMT3A*, *SF3B1*, *TP53* and *SRSF2*. While WES heterozygote genotypes for these variants will not include all variants with some degree of CHIP within these genes (as evidenced by many more individuals having non-zero alternate allele count at these locations, data not shown), it does capture CHIP-related somatic variants sufficiently to establish robust associations with longevity. In UKB the mean duration between the primary visit (blood draw date) and death is currently 9.2 years (± 3.8) and suggests that WES screening for CHIP variants may be used as a precision health tool to contribute to earlier cancer detection by assessing individuals with higher susceptibility risks. In addition to known cancer variants, such as breast cancer-related *BRCA1/BRCA2*, our study highlights novel associations that should be considered in cancer susceptibility screenings.

By combining large-scale GWAS with rare variant analysis, this study enhances our understanding of the genetic basis of human longevity. Our results emphasize the importance of understanding the genetic factors driving the most prevalent causes of mortality on a population level, highlighting the potential for early genetic testing to identify germline and somatic variants that place some individuals at risk of early death. Understanding the biological pathways through which these genes influence cancer and aging, as well as the environmental factors interacting with these pathways, will be essential for developing therapeutic targets aimed at extending healthy lifespan. Our study’s implications thus extend beyond genetics, as they touch on the broader aspects of health care, public health policy, and preventive strategies against age-related diseases.

In conclusion, this study enhances our understanding of the genetic basis of human longevity by combining large-scale GWAS with detailed rare variant analysis. The novel loci identified warrant further exploration to understand their biological roles and interactions with environmental factors, which will be crucial for unraveling the complex nature of aging and developing strategies to mitigate its adverse effects.

## Data Availability

Data supporting the findings of this study are available from the UK Biobank (UKB). Access to these data is available from the authors with UKB permission.

https://www.ukbiobank.ac.uk/enable-your-research/register

## Figures and Tables

**Supplementary Figure 1.**
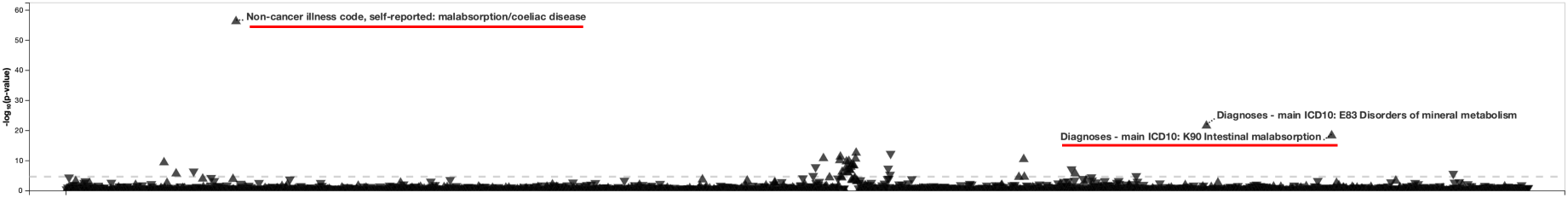
Phenome-wide association of rs13190937 on *ZSCAN23.* This analysis is based on PheWeb (https://pheweb.org/UKB-Neale/).

**Supplementary Figure 2.**
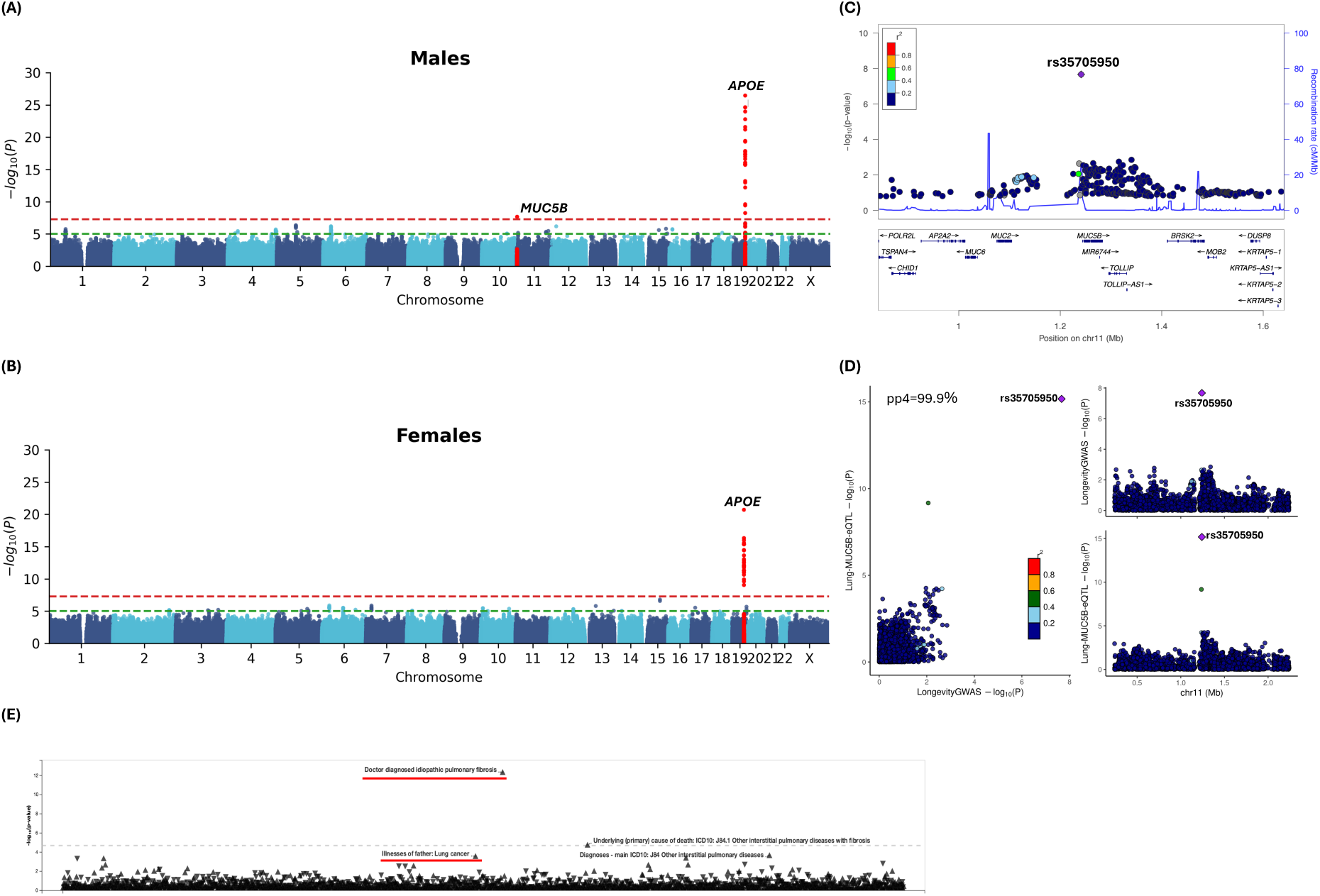
Sex-stratified common variant GWAS of longevity. Manhattan plot in males (A), and females (B). Locuszoom (C) and colocalization (D) plots at the *MUC5B* locus in males, colocalized with *MUC5B* eQTL in lung tissue in GTEx. PP4: posterior probability of colocalization. (E) Phenom-wide association of rs35705950. This analysis is based on PheWeb (https://pheweb.org/UKB-Neale/).

**Supplementary Figure 3.**
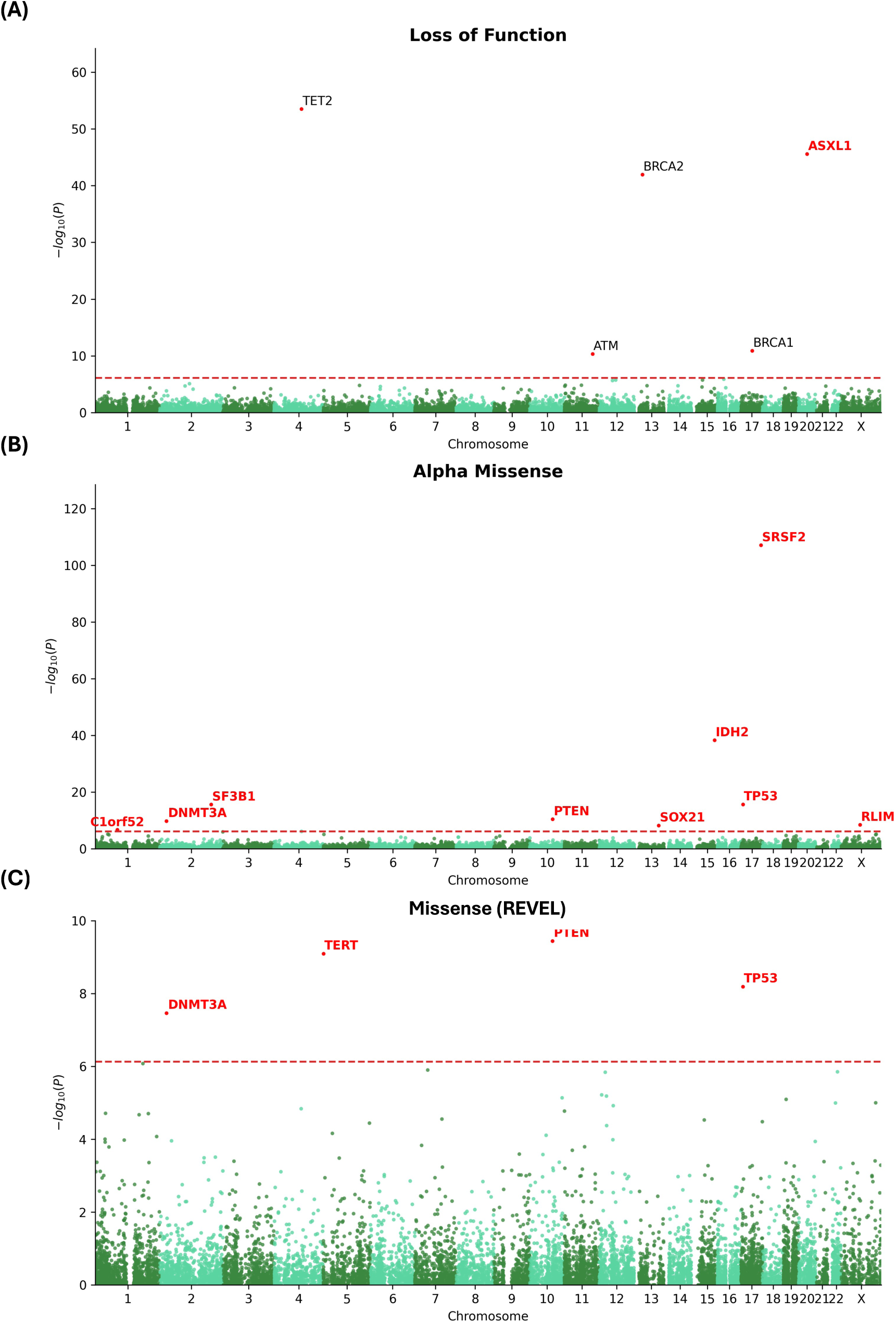
Rare variant SKAT-O association with longevity considering 3 categories: Loss-of-function (A), Alpha Missense (B), and REVEL (C). Novel genes are highlighted in red.

**Supplementary Figure 4.**
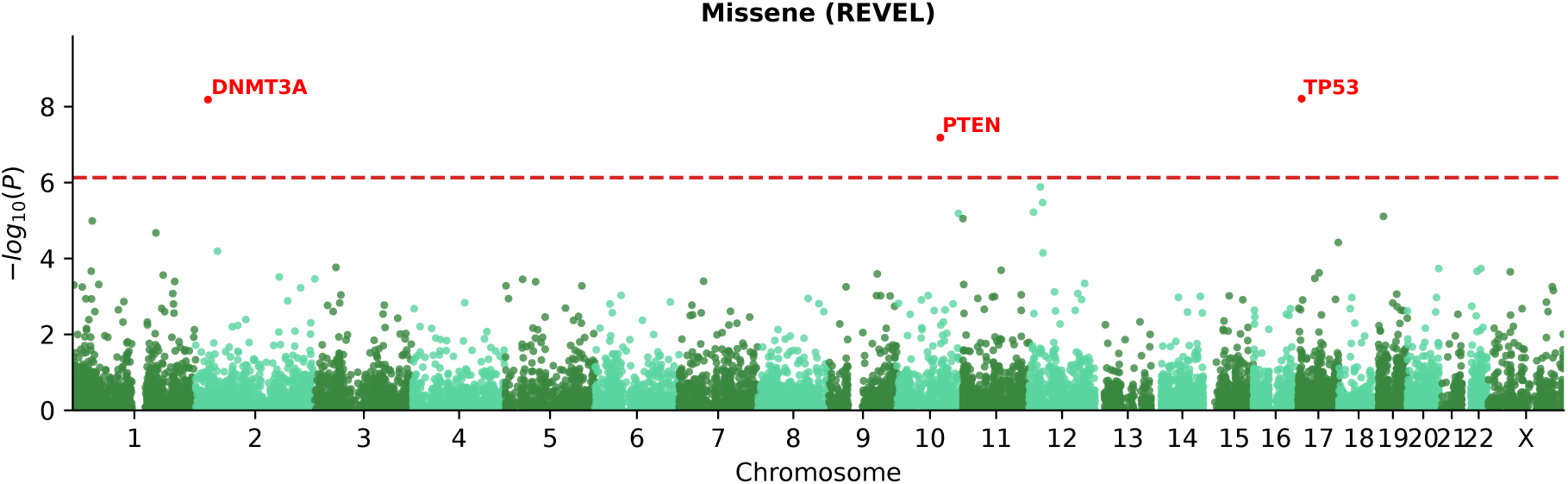
Rare variant burden association with longevity considering REVEL pathogenic missense variants. Novel genes are highlighted in red.

**Supplementary Figure 5.**
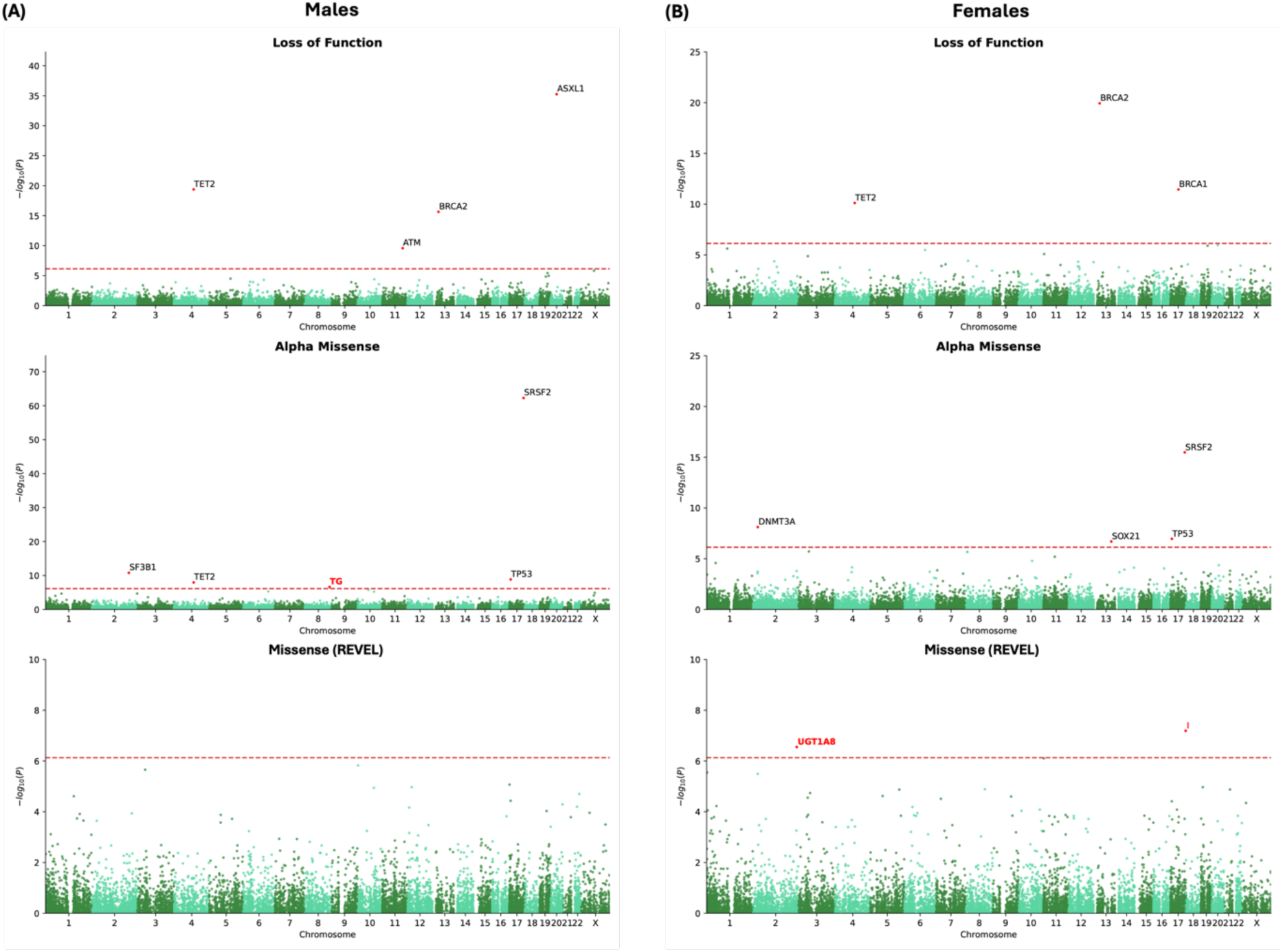
Sex-stratified rare variant burden association with longevity considering 3 categories for each sex: Loss-of-function (A), Alpha Missense (B), and REVEL (C). Novel genes are highlighted in red.

**Supplementary Figure 6.**
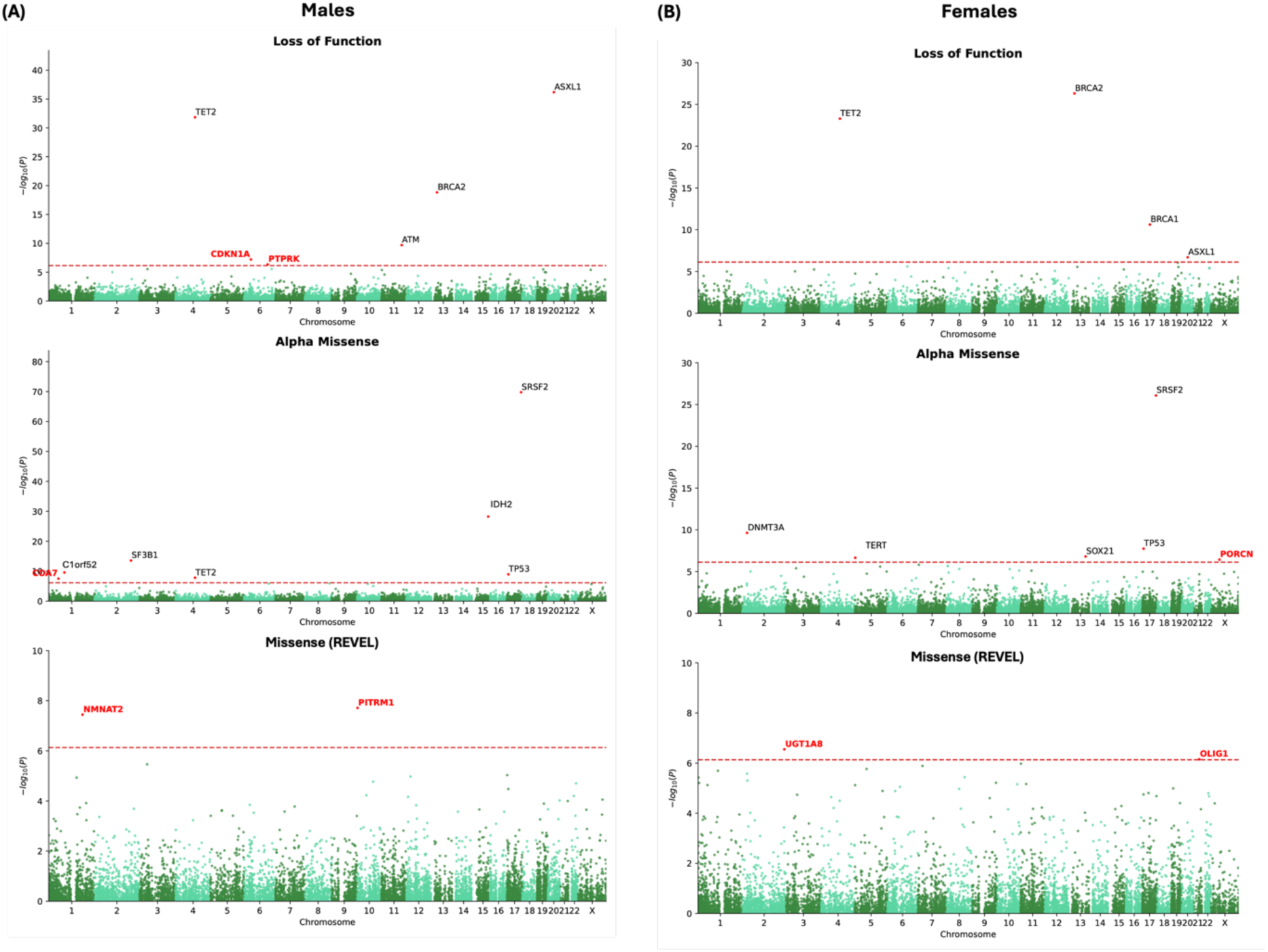
Sex-stratified rare variants SKAT-O association with longevity considering 3 categories for each sex: Loss-of-function (A), Alpha Missense (B), and REVEL (C). Novel genes are highlighted in red.

**Supplementary Figure 7.**
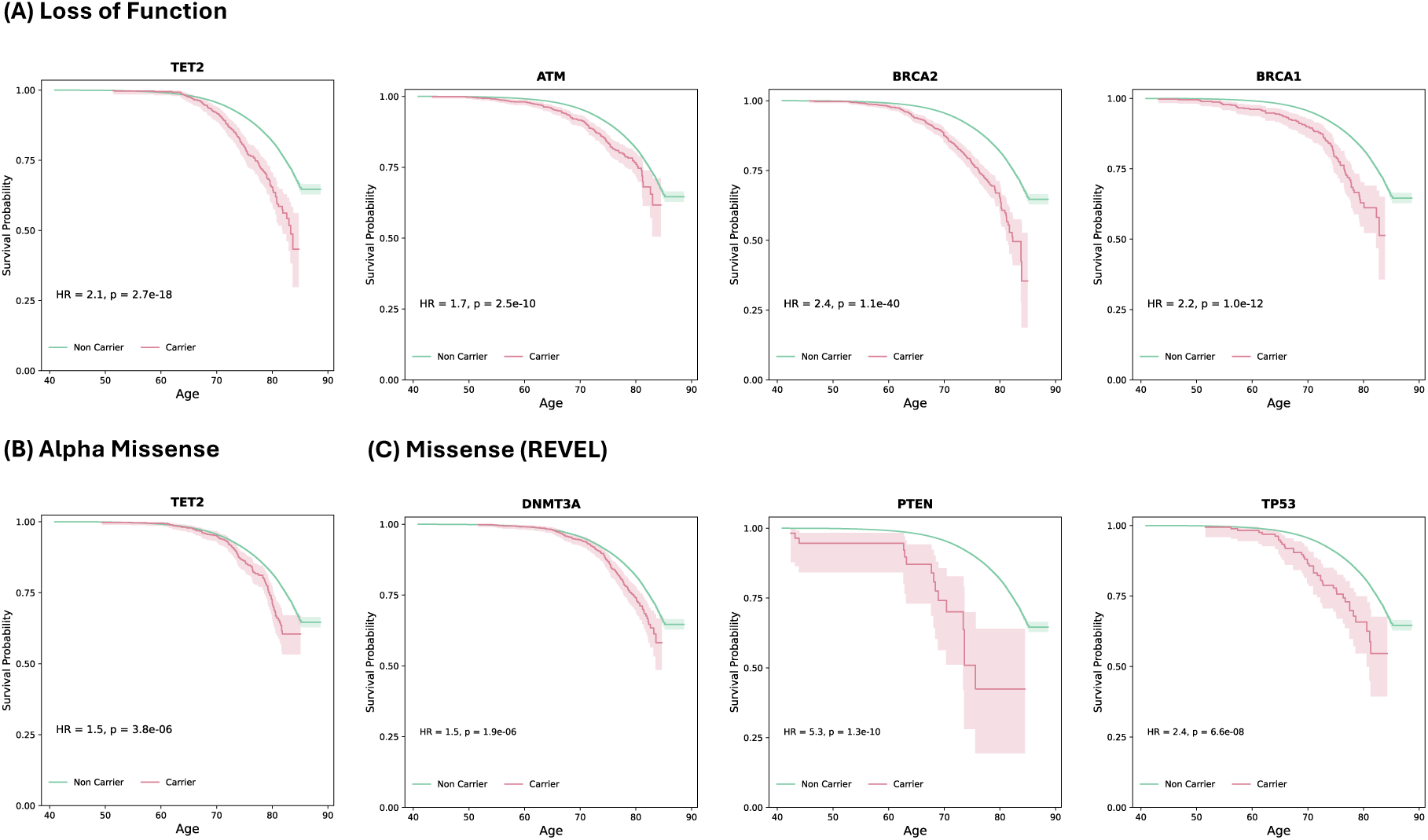
Survival curves comparing carriers and non-carriers of variants considered on genes with a significant burden of loss-of-function (*TET2*, *ATM*, *BRCA2* and *BRCA1*) (A), AlphaMissense pathogenic (B) variants (*TET2*), missense variants predicted by REVEL (*DNMT3A*, *PTEN* and *TP53*) (C)

**Supplementary Figure 8.**
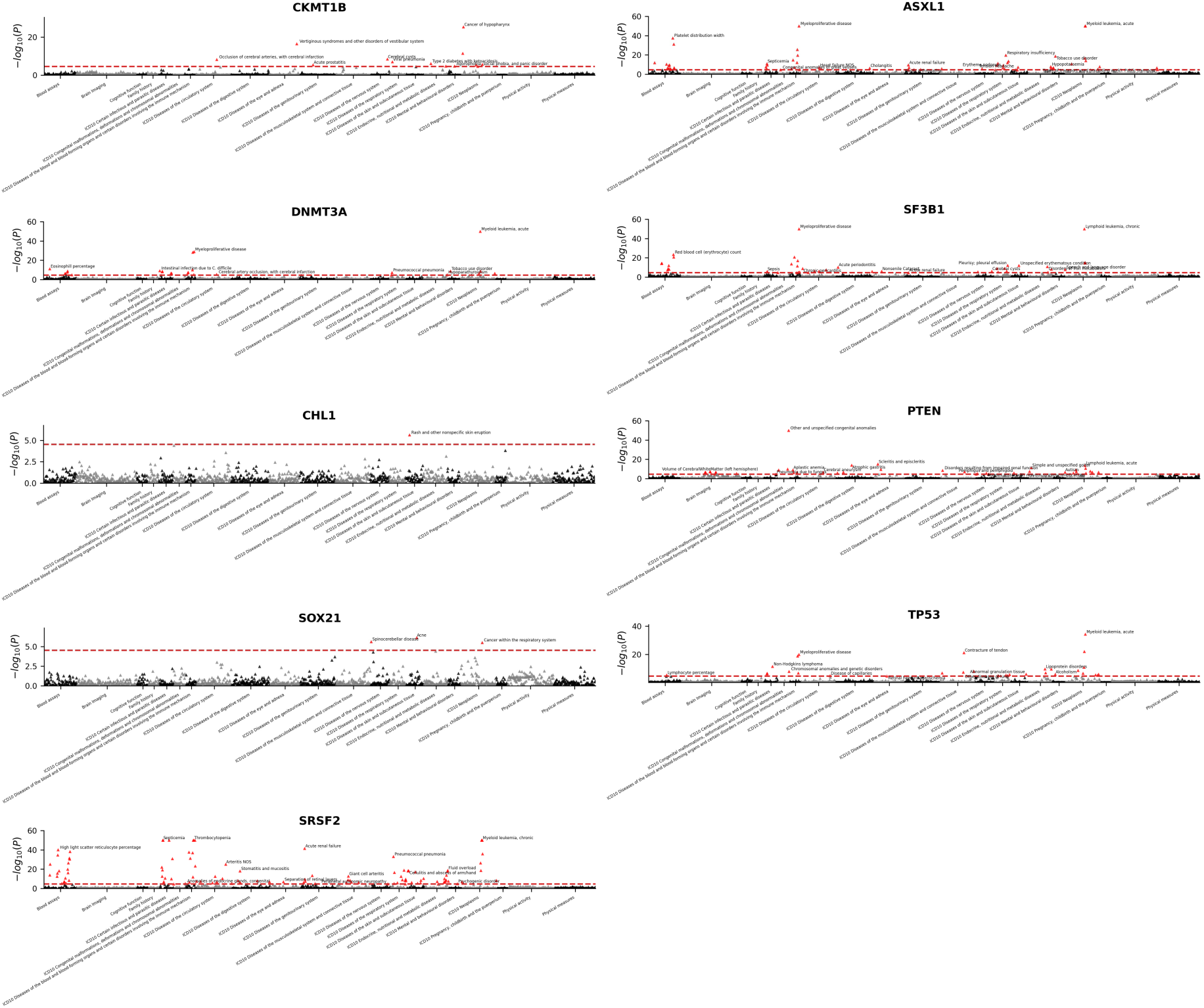
Phenome-wide association of the burden of rare variants at the nine novel genes identified in our burden test. Variants considered correspond to loss-of-function and Alpha missense defined variants. P-values less than 1.0×10^−50)^ are capped at 50.

**Supplementary Figure 9.**
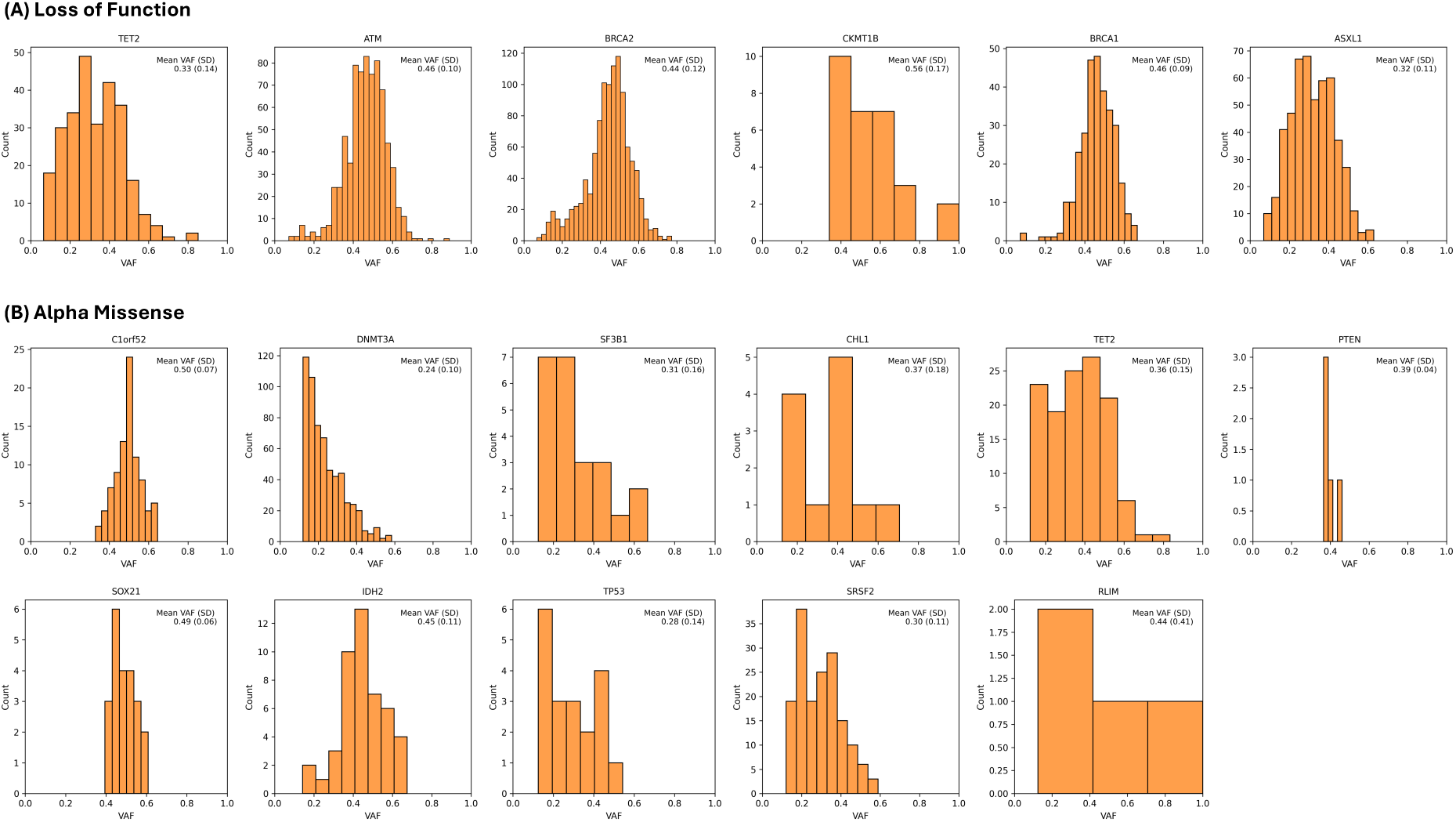
Variant allelic fraction distribution per gene for variants considered in each category: Loss-of-function (A) and Alpha Missense (B).

**Supplementary Table 1.** Demographics of European ancestry in the analyses.

|  | All |  |  | Male |  |  | Female |  |  |
| --- | --- | --- | --- | --- | --- | --- | --- | --- | --- |
|  | Total | Living | Deceased | Total | Living | Deceased | Total | Living | Deceased |
| N | 393,833 | 358,282 | 35,551 | 180,970 | 159,911 | 21,059 | 212,863 | 198,371 | 14,492 |
| Last known age | 70.8<br>± 7.9 | 70.7<br>± 8.0 | 71.2<br>± 7.5 | 70.9<br>± 8.0 | 70.8<br>± 8.1 | 71.3<br>± 7.4 | 70.7<br>± 7.9 | 70.7<br>± 7.9 | 71.1<br>± 7.6 |
| *APOE* ε4 carrier | 113,437<br>(28.8%) | 102,360<br>(28.6%) | 11,077<br>(31.2%) | 52,190<br>(28.8%) | 45,635<br>(28.5%) | 6,555<br>(31.1%) | 61,247<br>(28.8%) | 56,725<br>(28.6%) | 4,522<br>(31.2%) |

**Supplementary Table 2.** Significant genes for burden and SKAT-O association of rare variants, considering missense variants with REVEL > 75. Gene names in bold font represent novel associations in REVEL.

| Variant Class | Chr | Gene | # of variants | # of carriers | Burden<br>*p*-value | SKAT-O<br>*p*-value |
| --- | --- | --- | --- | --- | --- | --- |
| REVEL (>75) | 2 | *DNMT3A* | 116 | 831 | $6.6 \times 10^{-9}$ | $3.5 \times 10^{-8}$ |
| | 5 | ***TERT*** | 31 | 60 | $2.6 \times 10^{-3}$ | $8.1 \times 10^{-10}$ |
| | 10 | *PTEN* | 42 | 45 | $6.6 \times 10^{-8}$ | $3.6 \times 10^{-10}$ |
| | 17 | *TP53* | 47 | 173 | $6.2 \times 10^{-9}$ | $6.5 \times 10^{-9}$ |

**Supplementary Table 3.**
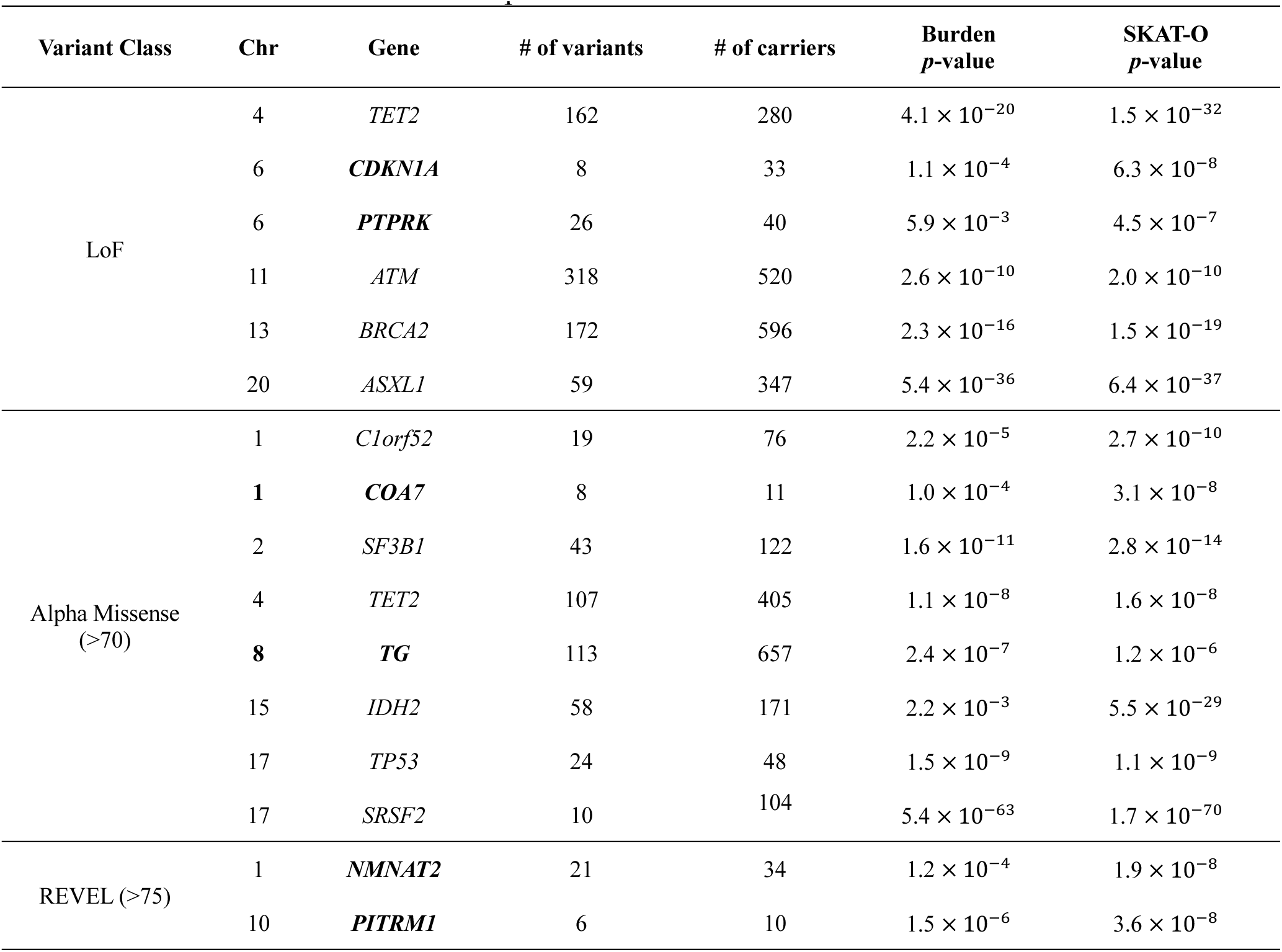
Significant genes for burden and SKAT-O association of rare variants in males. Genes in bold font represent novel associations in males.

**Supplementary Table 4.** Significant genes for burden and SKAT-O association of rare variants in females. Genes in bold font represent novel associations in females.

| Variant Class | Chr | Gene | # of variants | # of carriers | Burden<br>*p*-value | SKAT-O<br>*p*-value |
| --- | --- | --- | --- | --- | --- | --- |
| LoF | 4 | *TET2* | 151 | 283 | $7.8 \times 10^{-11}$ | $5.1 \times 10^{-24}$ |
| | 13 | *BRCA2* | 182 | 675 | $1.2 \times 10^{-20}$ | $4.8 \times 10^{-27}$ |
| | 17 | *BRCA1* | 80 | 217 | $3.6 \times 10^{-12}$ | $2.4 \times 10^{-11}$ |
| | 20 | *ASXL1* | 46 | 186 | $1.1 \times 10^{-6}$ | $2.0 \times 10^{-7}$ |
| Alpha Missense<br>(>70) | 2 | *DNMT3A* | 135 | 671 | $7.6 \times 10^{-9}$ | $2.3 \times 10^{-10}$ |
| | 5 | *TERT* | 22 | 27 | $1.7 \times 10^{-3}$ | $2.2 \times 10^{-7}$ |
| | 13 | *SOX21* | 34 | 251 | $2.0 \times 10^{-7}$ | $1.6 \times 10^{-7}$ |
| | 17 | *SRSF2* | 10 | 37 | $3.3 \times 10^{-16}$ | $8.4 \times 10^{-27}$ |
| | 17 | *TP53* | 23 | 52 | $1.1 \times 10^{-7}$ | $1.8 \times 10^{-8}$ |
| | X | ***PORCN*** | 11 | 32 | $5.6 \times 10^{-4}$ | $3.7 \times 10^{-7}$ |
| REVEL (>75) | 2 | ***UGT1A8*** | 2 | 18 | $2.8 \times 10^{-7}$ | $2.8 \times 10^{-7}$ |
| | 21 | ***OLIG1*** | 7 | 18 | $1.3 \times 10^{-5}$ | $7.0 \times 10^{-7}$ |

**Supplementary Table 5.** Lead variant association per gene among significant genes in the burden and SKAT-O tests. Only significant variant associations with at least 3 minor allele counts per gene are reported in this table.

| Variant Class | Chr | Gene | Variant | MA | MAC | AM | HR | *p*-value | Reported |
| --- | --- | --- | --- | --- | --- | --- | --- | --- | --- |
| REVEL (>75) | 2 | *DNMT3A* | rs367909007 | G | 14 | 0.983 | 4.3 | $1.2 \times 10^{-3}$ | - |
| | 5 | *TERT* | rs1043358053 | C | 5 | 0.926 | 11.9 | $7.4 \times 10^{-7}$ | - |
| | 10 | *PTEN* | rs587782350 | C | 3 | 0.941 | 20.4 | $2.6 \times 10^{-3}$ | - |
| | 17 | *TP53* | rs11540652 | T | 5 | 0.996 | 10.0 | $6.6 \times 10^{-5}$ | Gastric Cancer [70],<br>Ovarian Cancer [71] |

**Supplementary Table 6.** Mean variant allelic fraction per gene across participants included in the corresponding gene-level Burden/SKAT-O analysis.

| Variant Class | Chr | Gene | # of subjects | # of variants | Mean VAF (SD) |
| --- | --- | --- | --- | --- | --- |
| LoF | 4 | *TET2* | 266 | 133 | 0.33 (0.14) |
|  | 11 | *ATM* | 734 | 128 | 0.46 (0.10) |
|  | 13 | *BRCA2* | 1,061 | 162 | 0.44 (0.12) |
|  | 15 | *CKMT1B* | 29 | 8 | 0.56 (0.17) |
|  | 17 | *BRCA1* | 302 | 74 | 0.46 (0.09) |
|  | 20 | *ASXL1* | 502 | 30 | 0.32 (0.11) |
| Alpha Missense | 1 | *C1orf52* | 87 | 11 | 0.50 (0.07) |
|  | 2 | *DNMT3A* | 593 | 33 | 0.24 (0.10) |
|  | 2 | *SF3B1* | 23 | 5 | 0.31 (0.16) |
|  | 3 | *CHL1* | 12 | 3 | 0.37 (0.18) |
|  | 4 | *TET2* | 123 | 41 | 0.36 (0.15) |
|  | 10 | *PTEN* | 5 | 3 | 0.39 (0.04) |
|  | 13 | *SOX21* | 22 | 6 | 0.49 (0.06) |
|  | 15 | *IDH2* | 46 | 14 | 0.45 (0.11) |
|  | 17 | *TP53* | 19 | 7 | 0.28 (0.14) |
|  | 17 | *SRSF2* | 164 | 6 | 0.30 (0.11) |
|  | X | *RLIM* | 4 | 2 | 0.44 (0.41) |

## Acknowledgements

This research has been conducted using the UK Biobank Resource under application number 45420. We thank all the participants and researchers of UK Biobank for making these data open and accessible to the research community.

## Authors’ contributions

J.P conducted all the analyses, prepared all figures and wrote the manuscript. A.P.T contributed to the manuscript writing. L.T provided critical comment on the manuscript. M.D.G and Y.L.G planned, organized and supervised the entire study and revised the manuscript. All authors have approved the submitted version.

## Data availability

GWAS summary statistics for this study are available in the GWAS Catalog. Data supporting the findings of this study are available from the UK Biobank (UKB). Access to these data is available from the authors with UKB permission

## Code availability

The codes used for analyses in the present study are available at the following link: https://github.com/Junkkkk/Lifespan-studies

## Consent for publication

Not applicable.

